# TLR7 Variants Enhance Responsiveness and Broaden RNA Specificity To Drive Human Lupus

**DOI:** 10.1101/2025.11.07.25338211

**Authors:** Asuka Takayama, Kazuyuki Meguro, Takuma Shibata, Fumiya Yamaide, Yuji Imai, Yoshiyuki Goto, Yosuke Nishio, Taichi Oso, Tomoyuki Numata, Kotaro Suzuki, Keishi Etori, Takahiro Kageyama, Takashi Ito, Ai Maeda, Hiroyuki Yagyu, Takeshi Yamamoto, Junichi Ishikawa, Kiyoshi Hirahara, Hiromichi Hamada, Tomoo Ogi, Tomohiko Ichikawa, Emi K. Nishimura, Tsuyoshi Koide, Toshiyuki Shimizu, Kensuke Miyake, Hiroshi Nakajima

**Affiliations:** Department of Allergy and Clinical Immunology, Graduate School of Medicine, Chiba University, Chiba, Japan; Clinical Genetics, Chiba University Hospital, Chiba, Japan; Chiba University Synergy Institute for Futuristic Mucosal Vaccine Research and Development (cSIMVa), Chiba University, Chiba, Japan; Division of Aging and Regeneration, The Institute of Medical Science, The University of Tokyo, Tokyo, Japan; Department of Pediatrics, Graduate School of Medicine, Chiba University, Chiba, Japan; Mouse Genomics Resource Laboratory, National Institute of Genetics, Mishima, Japan; Division of Molecular Immunology, Medical Mycology Research Center, Chiba University, Chiba, Japan; Division of Pandemic and Post-disaster Infectious Diseases, Research Institute of Disaster Medicine, Chiba University, Chiba, Japan; Division of Infectious Disease Vaccine R&D, Research Institute of Disaster Medicine, Chiba University, Chiba, Japan; Department of Genetics, Research Institute of Environmental Medicine, Nagoya University, Nagoya, Japan; Department of Human Genetics and Molecular Biology, Graduate School of Medicine, Nagoya University, Nagoya, Japan; Medical Genomics Center, Nagoya University Hospital, Nagoya, Japan; Department of Bioscience and Biotechnology, Graduate School of Bioresource and Bioenvironmental Sciences, Kyushu University, Fukuoka, Japan; Department of Immunology, Graduate School of Medicine, Chiba University, Chiba, Japan; Division of Animal Medical Science, Center for One Medicine Innovative Translational Research, Nagoya University, Nagoya, Japan; Division of Molecular Physiology and Dynamics, Institute for Glyco-core Research, Tokai National Higher Education and Research System, Nagoya, Japan; Laboratory of Protein Structural Biology, Graduate School of Pharmaceutical Sciences, The University of Tokyo, Tokyo, Japan

## Abstract

Dysregulated nucleic acid sensing underlies lethal viral infections and autoimmunity. Endosomal Toll-like receptor 7 (TLR7) recognizes guanosine and oligoribonucleotides (ORNs) from single-stranded RNA via two ligand-binding sites in its leucine-rich repeat (LRR) ectodomain. Recent genetic studies have demonstrated that *TLR7* gain-of-function (GOF) variants cause childhood lupus or immune thrombocytopenic purpura by enhancing TLR7 responses to canonical TLR7 ligands. However, no study has demonstrated that altered ligand specificity of TLR7 contributes to autoimmunity. Here, we provide direct evidence that relaxation of ligand specificity by *TLR7* genetic variants causes lupus in both humans and mice. We describe a pediatric systemic lupus erythematosus (SLE) patient carrying a private GOF *TLR7* missense variant, V825M (VarM), located within the C-terminal region of the LRR (LRR-CT). Knock-in mice harboring *VarM* allele spontaneously develop lupus-like systemic inflammation, including duodenitis, recapitulating the patient’s clinical features. Mechanistically, the VarM variant markedly amplifies NF-κB activation by synthetic small-molecule agonists and guanosine analogs. Notably, the VarM variant broadens the spectrum of ORN sequences that can engage the receptor. Moreover, certain pathogenic *TLR7* variants located at the dimerization interface display functional properties closely resembling those of VarM. Collectively, our findings establish a new paradigm in which alterations in ligand recognition specificity by *TLR7* genetic variations can trigger human autoimmune disease.

## Introduction

Nucleic acid sensing receptors constitute a first line of defense against viruses, yet their dysregulation provokes life-threatening viral infections and autoimmunity (*1–3*). Among them, Toll-like receptor 7 (TLR7) resides in endolysosomal compartments and recognizes single-stranded RNA (ssRNA) from viruses, such as SARS-CoV-2, as well as endogenous ssRNA released by dying host cells. Upon ligand engagement, the signaling adaptor MyD88 is recruited to the cytoplasmic Toll/IL-1 receptor (TIR) domain, thereby activating nuclear factor κB (NF-κB) and interferon regulatory factor 7 (IRF7) pathways to induce proinflammatory cytokines and type I interferons, respectively. Recent structural and biochemical studies have shown that TLR7 does not directly bind intact ssRNA. Instead, it senses RNA degradation products, including guanosine, 2′, 3′-cGMP, and short oligoribonucleotides (ORNs). Among ORNs, TLR7 preferentially recognizes those containing consecutive uridines (2U-RNA) over those with a single uridine (1U-RNA) (*4–6*). Thus, TLR7 functions as a metabolic sensor of lysosomal RNA catabolism.

Animal models highlight the pathological potential of TLR7 in autoimmune and autoinflammatory diseases, particularly systemic lupus erythematosus (SLE), a chronic autoimmune disorder characterized by multi-organ damage and the presence of anti-nuclear antibody (ANA). TLR7 transgenic mice and BXSB-Yaa male mice, which carry an extra *Tlr7* copy, spontaneously develop fatal lupus-like glomerulonephritis (*7, 8*), whereas *Tlr7* deletion in MRL/lpr mice, another SLE model, ameliorates disease severity and reduces ANA titers (*9–11*). Consistently, whole-genome sequencing in pediatric SLE patients has uncovered gain-of-function (GOF) variants in TLR7 and its chaperone protein UNC93B1 that define monogenic forms of SLE (*12–17*). These findings indicate that tight control of TLR7-UNC93B1 axis is indispensable for maintaining immune tolerance in humans. However, in previous studies of SLE, analyses of TLR7 have been limited to responsiveness to canonical ligands, whereas the potential impact of patient-derived TLR7 variants on the spectrum of ligand recognition has remained unexplored. Here, we identify a novel TLR7 missense variant from a patient with childhood-onset lupus and demonstrate that a group of TLR7 GOF variants, including our novel variant, broaden the spectrum of ligand RNA sequences, rather than merely enhancing responses to canonical ligands.

## Results

### Identification of a novel heterozygous variant in *TLR7* from a patient with severe lupus

We identified a patient with severe childhood lupus complicated by neurodevelopmental problems (Supplementary Text). The patient exhibited failure to thrive and neurodevelopmental delay after birth. During the first few years of life, she developed chronic immune thrombocytopenia. Several years later, she presented with a fever, nephritis, and duodenitis (Fig. 1A). She also had elevated autoantibodies, fulfilling the diagnostic criteria for SLE (Supplementary Text). Genomic analysis, including whole-exome sequencing (WES), revealed a *de novo* heterozygous missense variant in *TLR7*, V825M, which had not been previously reported (Fig. 1, B and C). The patient also carried a *de novo* heterozygous nonsense variant in *SOX5* (R203X; fig. S1A), which accounts for the neurodevelopmental symptoms but not the immunological phenotypes (*18*). Neither homozygotes nor heterozygotes of *TLR7* V825M were found in the Genome Aggregation Database (gnomAD), and the variant has a high Combined Annotation Dependent Depletion (CADD) score (*18*), comparable to previously reported pathogenic variants of *TLR7* (*12*) (fig. S1B). WES revealed no other variants in the patient known to cause inborn errors of immunity. These findings strongly suggest that the immune abnormalities observed in this SLE patient are associated with the *TLR7* V825M variant.

**Figure 1.**
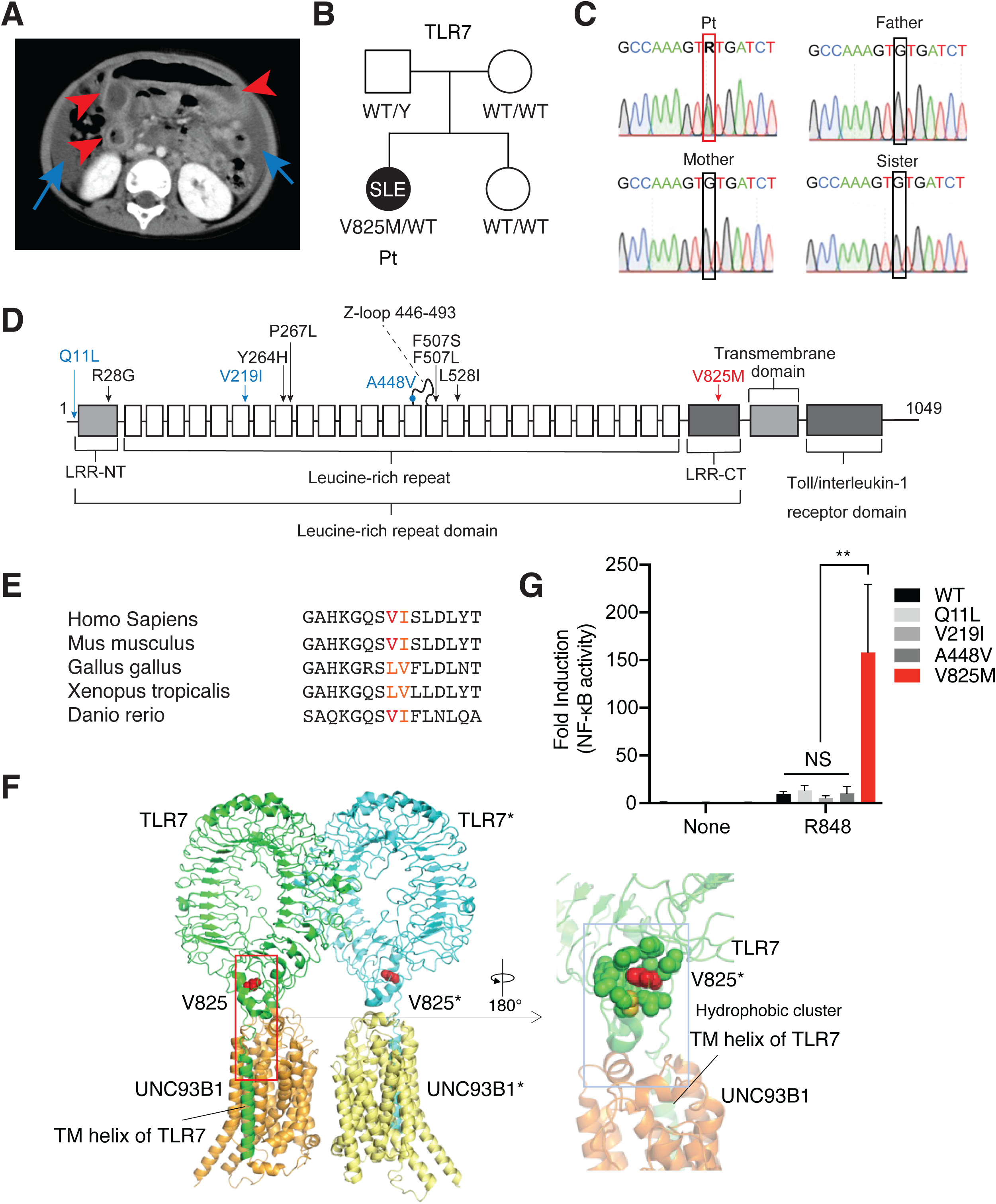
A novel missense variant in the LRR-CT domain of TLR7 associated with severe childhood lupus. (**A**) Computed tomography showing the enteritis and peritonitis in the patient. Red arrowhead, thickness and dilation of the intestinal wall with fluid accumulation; blue arrow, ascites. (**B**) Patient pedigree and TLR7 genotyping. Filled symbol, affected; open symbols, unaffected. (**C**) Chromatograms of Sanger sequencing of gDNA from the patient and family for TLR7 V825M variant. R indicates A or G. (**D**) Schema of TLR7 and the location of TLR7 variants. LRR-NT, N-terminal leucine-rich repeat; LRR-CT, C-terminal leucine-rich repeat. Blue, benign variants from gnomAD; Black, GOF variants previously reported; Red, variant identified in this study. (**E**) Amino acid conservation across five species. Red, conserved valine; orange, hydrophobic amino acids around the variant. **(F**) Structure of human TLR7 complexed with UNC93B1. (Left) Overall structure of TLR7 in complex with UNC93B1 (PDB ID: 7CYN). TLR7 and UNC93B1 are shown in ribbon diagrams, and Val825 of TLR7 is represented as a red sphere. The TLR7 molecules are colored green and light blue, and UNC93B1 molecules are colored brown and yellow. V825 is colored red. (Right) Close-up view of the interface between TLR7 and UNC93B1. Hydrophobic cluster close to Val825 is shown as spheres in green. (**G**) NF-κB dependent luciferase reporter assay of human TLR7 variants in HEK 293T cells co-expressing TLR7 and UNC93B1. WT, wild type; V825M, variant identified in this study; Q11L, V219I, A448V, benign variants from gnomAD. Cells were unstimulated or stimulated with R848. Data represent the mean fold induction of NF-κB activity ± SD, calculated as the RLU of stimulated cells divided by the RLU of non-stimulated cells. The data shown are representative of more than three independent experiments. Statistical analysis was performed using one-way ANOVA. *, p<0.05; **, p<0.01; ***, p<0.001; NS, not significant.

Structurally, TLR7 comprises three domains: a leucine-rich repeat (LRR) ectodomain that binds ligands, a single transmembrane helix (TM), and a cytoplasmic Toll/IL-1 receptor (TIR) domain that transduces signals. Unlike other SLE-associated *TLR7* variants located in the LRR region, such as Y264H, P267L, F507L/S, and L528I, V825M resides in the C-terminal non-canonical LRR (LRR-CT), and the amino acid sequence surrounding V825 in TLR7 is highly conserved across vertebrates (Fig. 1, D and E). The structure shows that V825 forms part of a hydrophobic cluster positioned immediately above the TM helix (Fig. 1F). Notably, the residues constituting this hydrophobic cluster are conserved among other TLR family members (*19*) (fig. S1C). To investigate the activity of the TLR7 V825M variant, we performed an NF-κB-dependent luciferase reporter assay. We also investigated the activity of benign variants from the general population (Q11L, V219I, A448V). We observed no activation of NF-κB for the V825M as well as WT and other benign variants (Fig. 1G). Interestingly, V825M showed strong activity compared to WT and benign variants in response to small chemical TLR7 ligand R848 (Fig. 1G). Taken together, we identified a novel TLR7 missense variant which has the potential to explain the severe childhood lupus.

### VarM mice develop spontaneous lupus-like autoimmune diseases

V825 of human TLR7 is conserved in mouse Tlr7 as V826 (Fig. 1E), and the *Tlr7* V826M allele confers a gain-of-function activity comparable to the human V825M variant in response to R848 (fig. S2A), a synthetic small-molecule ligand. To assess whether the SLE patient’s clinical features were attributable to *TLR7* V825M rather than the *SOX5* R203X variant, we generated *Tlr7* V826M knock-in mice on a C57BL/6 background using CRISPR-Cas9 editing (fig. S2B). We designated *Tlr7* V826M as VarM and named the knock-in strain VarM mice. Because the *Tlr7* gene is X-linked, pathology driven by the *VarM* allele follows an X-linked dominant inheritance pattern. The overall survival rate of VarM mice was lower than that of wild-type (WT) mice (Fig. 2A) and hemizygous VarM male mice showed reduced body weight compared with WT controls (fig. S2C). Heterozygous and homozygous female VarM mice developed marked splenomegaly (Fig. 2B and fig. S2D), and the normal red and white pulp architecture was profoundly disrupted (Fig. 2C). Histological examination revealed spontaneous inflammation in the liver and kidney in VarM mice (Fig. 2, D – F and fig. S2E), resembling lesions observed in the *Tlr7* Y264H lupus model (12). Consistent with chronic inflammation and liver injury, serum albumin levels were reduced in VarM mice, whereas serum aspartate aminotransferase, alanine aminotransferase, and lactate dehydrogenase levels were elevated (fig. S3, A – D). Serum total bilirubin levels were significantly increased in VarM mice (fig. S3E). Serum creatinine levels were slightly elevated in VarM mice, but the increase was not statistically significant (fig. S3F). High-density lipoprotein cholesterol and total cholesterol levels were decreased in VarM mice (fig. S3, G and H), consistent with interferon-driven reprogramming of lipid metabolism (*20*). Strikingly, VarM mice also developed spontaneous duodenitis, an intestinal manifestation not previously described in other TLR7-dependent lupus models, yet consistent with the clinical presentation of our patient (Fig. 2G). In addition, VarM mice exhibited anemia (fig. S2F) and thrombocytopenia (Fig. 2H, fig. S2G), and produced antinuclear antibodies (ANAs) in almost all mice harboring the *VarM* allele (Fig. 2I and J), which are typical clinical features of lupus patients. We generated two independent VarM strains from different founders and obtained consistent results across both strains (data not shown). Collectively, these data establish VarM mice as a robust spontaneous lupus model that recapitulates the patient’s systemic inflammation, including unique intestinal involvement.

**Figure 2.**
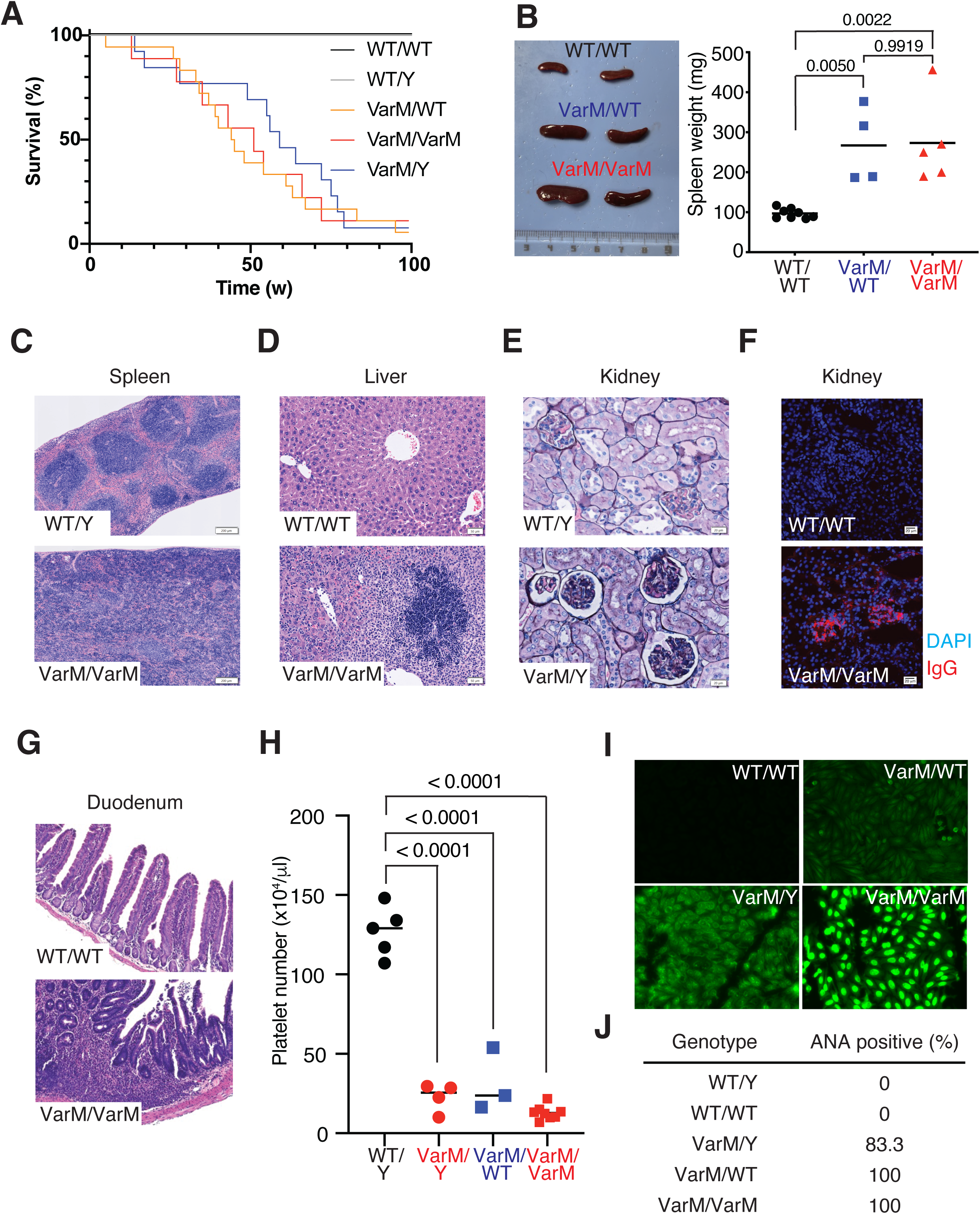
VarM mice develop lupus-like autoimmune disease. (**A)** The survival of VarM mice. n=15 (*Tlr7*^WT/WT^), n= 9 (*Tlr7*^WT/Y^), n=56 (*Tlr7*^VarM/WT^), n=10 (*Tlr7*^VarM/VarM^), n=28 (*Tlr7*^VarM/Y^). *Tlr*^WT/WT^ and *Tlr7*^WT/Y^ curves (both are 100 % survival) were graphically offset to improve visibility. (**B**) Weight of spleen from 20-week-old mice with the indicated genotypes. Data indicate mean, and statistical analyses were performed by one-way ANOVA. *, p<0.05; **, p<0.01; NS, not significant. (**C-G**) Histological analysis of organs affected in VarM mice. Hematoxylin-eosin (HE) stain of spleen (C) and liver (D). Periodic acid-methenamine silver (PAM) stain of kidney (E). Immunofluorescent staining showing C3 and IgG deposition in kidney (F). HE stain of duodenum (G). Scale bar, 100 μm. (**H**) Platelet count in peripheral blood from indicated mice at the age of 52 weeks. Data indicate mean of n=5 (WT/Y), n=4 (VarM/Y), n=3 (VarM/WT), and n=5 (VarM/VarM). Statistical analyses were performed using one-way ANOVA. ***, p<0.001. (**I**) Immunofluorescent microscopy of Hep-2 cells using serum from 12-week-old WT/WT (upper left), VarM/WT (upper right), VarM/Y (lower left), and VarM/VarM (lower right). (**J**) Proportion of ANA positive mice at 12-week-old. WT/WT, n=6; WT/Y, n=6; VarM/WT, n=6; VarM/Y, n=6; VarM/VarM, n=6.

### Immune cells from VarM mice show enhanced TLR7 responses

To investigate TLR7-driven immune changes in VarM mice, we analyzed splenocytes from VarM mice showing splenomegaly using multiparameter flow cytometry. Ly6C high classical and Ly6C low patrolling monocytes were both significantly increased in VarM mice (Fig. 3, A and B). While the frequency of B cells was unchanged (Fig. 3, C and D), CD4+ and CD8+ T cells were markedly reduced in VarM mice (Fig. 3E), mirroring phenotypes reported in another mouse model with enhanced TLR7 responses in macrophages (23). Notably, CD4/8 double negative (DN) T cells were expanded in VarM mice (Fig. 3F) and in the patient (Supplementary Text), reminiscent of DN T cell accumulation in autoimmune lymphoproliferative syndrome (ALPS) (24).

**Figure 3.**
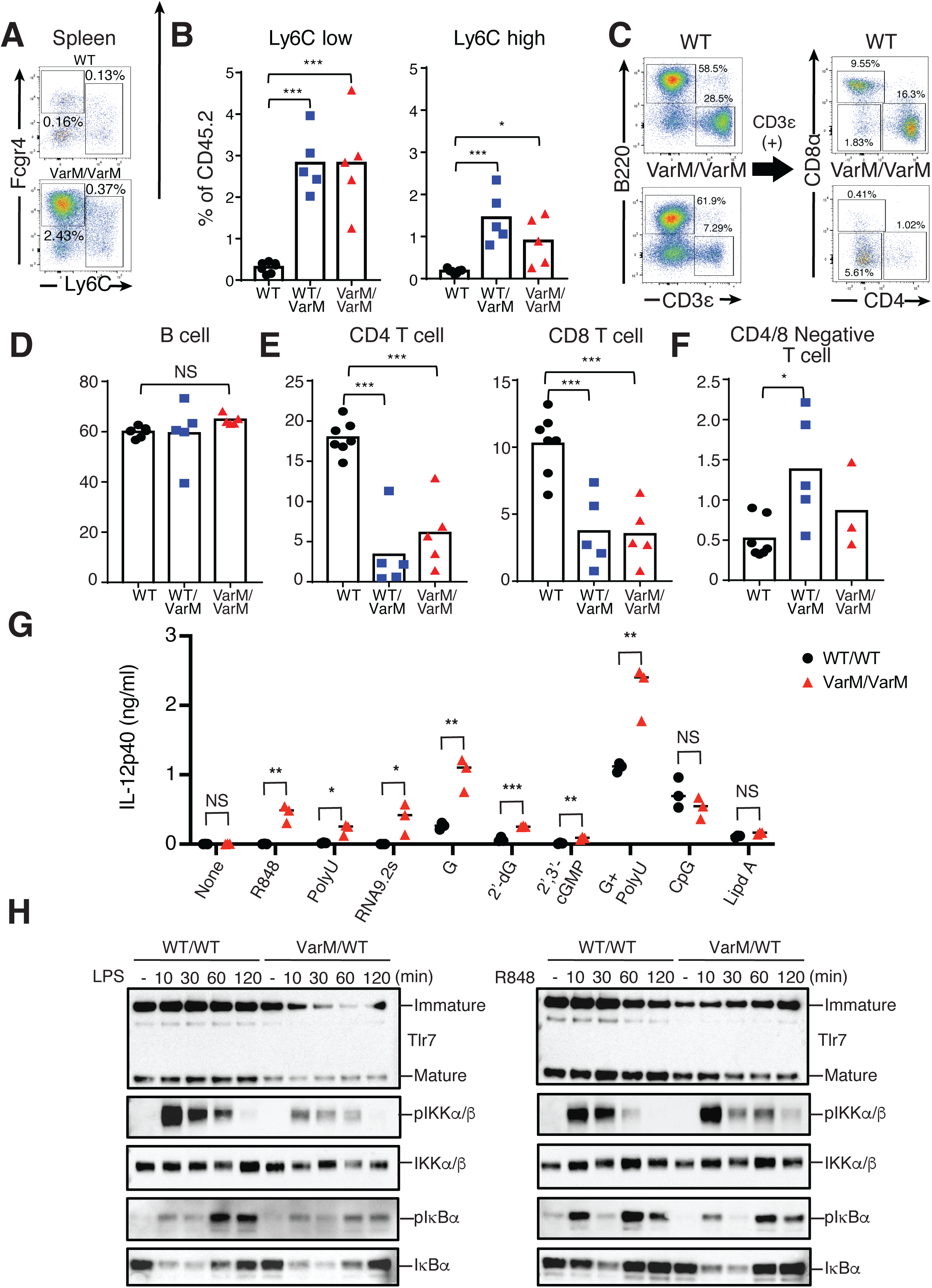
Immune cells from VarM mice exhibit hyperresponsiveness for TLR7 ligands. (**A**-**F)** Representative flow cytometry plots and the number of indicated immune cells in WT/WT, VarM/WT, and VarM/VarM mice. Data indicate mean. WT, n=7; VarM/WT, n=5; VarM/VarM, n= 3. Statistical analysis was performed using one-way ANOVA. *, p<0.05; **, p<0.01; ***, p<0.001; NS, not significant. (**G**) IL-12p40 production form BMDMs from WT/WT (n=3), VarM/WT (n=3), VarM/VarM (n=3) mice in response to low dose TLR7 ligands. G, guanosine. Data indicate mean and statistical analysis was performed using multiple unpaired t-test. *, p<0.05; **, p<0.01; ***, p<0.001; NS, not significant. (**H**) Immunoblot of NF-κB signaling molecules using BMDMs from VarM mice in response to LPS (left) and R848 (right).

We next evaluated TLR7 responses in VarM mice by stimulating bone marrow-derived macrophages (BMDMs). As expected from the NF-κB-dependent luciferase reporter assay (fig. S2A), robust production of pro-inflammatory cytokines was noted in BMDMs from *VarM/VarM* mice in response to a wide variety of canonical TLR7 ligands (Fig. 3G). Low doses of small chemical TLR7 ligand more clearly revealed the increased proinflammatory cytokine production in VarM derived BMDMs compared with WT cells (fig. S3, I and J). Interestingly, the *VarM* allele selectively enhanced responsiveness to lower, but not higher, ligand concentrations (fig. S3, I and J), indicating that V826M increases TLR7 sensitivity.

Consistently, a low dose of R848 induced more prolonged phosphorylation of IKKα/β (pIKKα/β) and degradation of IκBα in BMDMs from VarM mice, whereas LPS responses were unaffected (Fig. 3H). In summary, these data demonstrate that the V826M variant amplifies TLR7 signaling and lowers its activation threshold in both *in vivo* and *in vitro* settings.

### *TLR7* V825M variant relaxes ligand specificity and exaggerates TLR7 signaling

Our prior analyses of single–amino acid substitutions in the ligand-binding pockets of TLR8, another ssRNA sensor in the TLR7 family, revealed that such alterations can profoundly enhance receptor responsiveness to both small-molecule and RNA ligands. Notably, artificial mutations at the first binding site enabled TLR8, normally a uridine sensor, to recognize guanosine (*6, 21*). These observations suggest that even subtle amino acid substitutions can not only lower the signaling threshold but also broaden the ligand specificity of nucleic acid–sensing TLRs, potentially explaining the pathogenic effect of the VarM variant in SLE, although such dysregulation in ligand recognition of immune sensors has yet to be demonstrated in human inborn errors of immunity (IEIs). To test this hypothesis, we performed NF-κB reporter assays comparing V825M with other SLE-associated and benign TLR7 variants.

The V825M substitution resides in the LRR-CT of TLR7, whereas all previously described GOF variants found in SLE patients - R28G, Y264H, P267L, F507L, F507S and L528I - map to the LRR-NT or the canonical LRR scaffold (*12, 22–24*). R28G is located within LRR-NT, Y264H and P267L flank the guanosine-binding pocket, and F507L, F507S, and L528I cluster at the LRR dimerization interface (Fig. 4A). To compare their functional impact, we measured NF-κB-driven NanoLuc activity in HEK293T cells transfected with WT *TLR7*, the seven GOF variants, or benign *TLR7* variants identified in the general population (Q11L, V219I, A448V). None of the variants, including V825M, showed constitutive activity, indicating that ligand engagement is required for activation of GOF variants (Fig. 4B). After stimulation with the synthetic agonists R848 or CL264, V825M elicited the highest signal, exceeding both WT and all other variants (Fig. 4C). Dimerization-interface variants (F507L, F507S, L528I) showed a milder GOF phenotype with R848 compared to V825M and no enhanced response with CL264 (Fig. 4C). Because TLR7 principally senses guanosine (*4*), we next profiled responses to five nucleosides. Although WT TLR7 requires ORNs to sense guanosine analogs, V825M and the three dimerization-interface variants selectively amplified signaling to guanosine alone across a wide concentration range, but not to adenosine, uridine, cytidine or inosine (Fig. 4, D and E). Consistent results were obtained with guanosine analogs such as 2’-deoxyguanosine and 2′, 3′-cGMP, where V825M and dimerization-interface variants conferred the strongest NF-κB activation (Fig. 4, F and G), indicating that these SLE-associated variants can substantially lower the activation threshold for guanosine analog recognition. Because TLR7 also recognizes oligoribonucleotides, we examined variant responses to various ssRNA sequences (Fig. 5A). V825M markedly increased sensitivity to low doses of PolyU and RNA9.2s, known TLR7 ligands (Fig. 5B). Notably, V825M even responded to ssRNA40, a canonical TLR8 agonist, indicating altered sequence specificity of ssRNA recognition (Fig. 5B). We previously defined the sequence preferences of the ORN ligand-binding pocket in WT TLR7 (*4*). In the presence of guanosine, ssRNAs containing two or more consecutive uridines (2U-RNAs) elicit stronger TLR7 signaling than ssRNAs without uridine repeat (1U-RNAs). Moreover, TLR7 is most efficiently activated when a pyrimidine base (C or U), rather than a purine base (A or G), extends toward the 3’ end of the UU motif. V825M and the dimerization-interface variants responded robustly to UUC and UUG at concentrations that failed to activate WT TLR7 (Fig. 5C). When sub-threshold amounts of 2’, 3’-cGMP were added, which alone barely induced NF-κB activation, these variants further amplified NF-κB signaling in response to UUU, UUC, UUG and UUA ORNs (Fig. 5D). Thus, these variants relax the requirement for a pyrimidine residue at the 3’ end of 2U-RNAs, enabling recognition of a broader range of ssRNAs compared to WT TLR7. By contrast, the SLE-associated Y264H variant enhanced signaling only to UUG, indicating a gain-of-function mechanism distinct from that of V825M and dimerization-interface variants. Collectively, these series of reporter assays demonstrate that the *TLR7* V825M variant, together with a cluster of dimerization interface variants, broadens the spectrum of RNA sequences recognized by TLR7, thereby driving pathological hyperresponsiveness that contributes to SLE onset.

**Figure 4.**
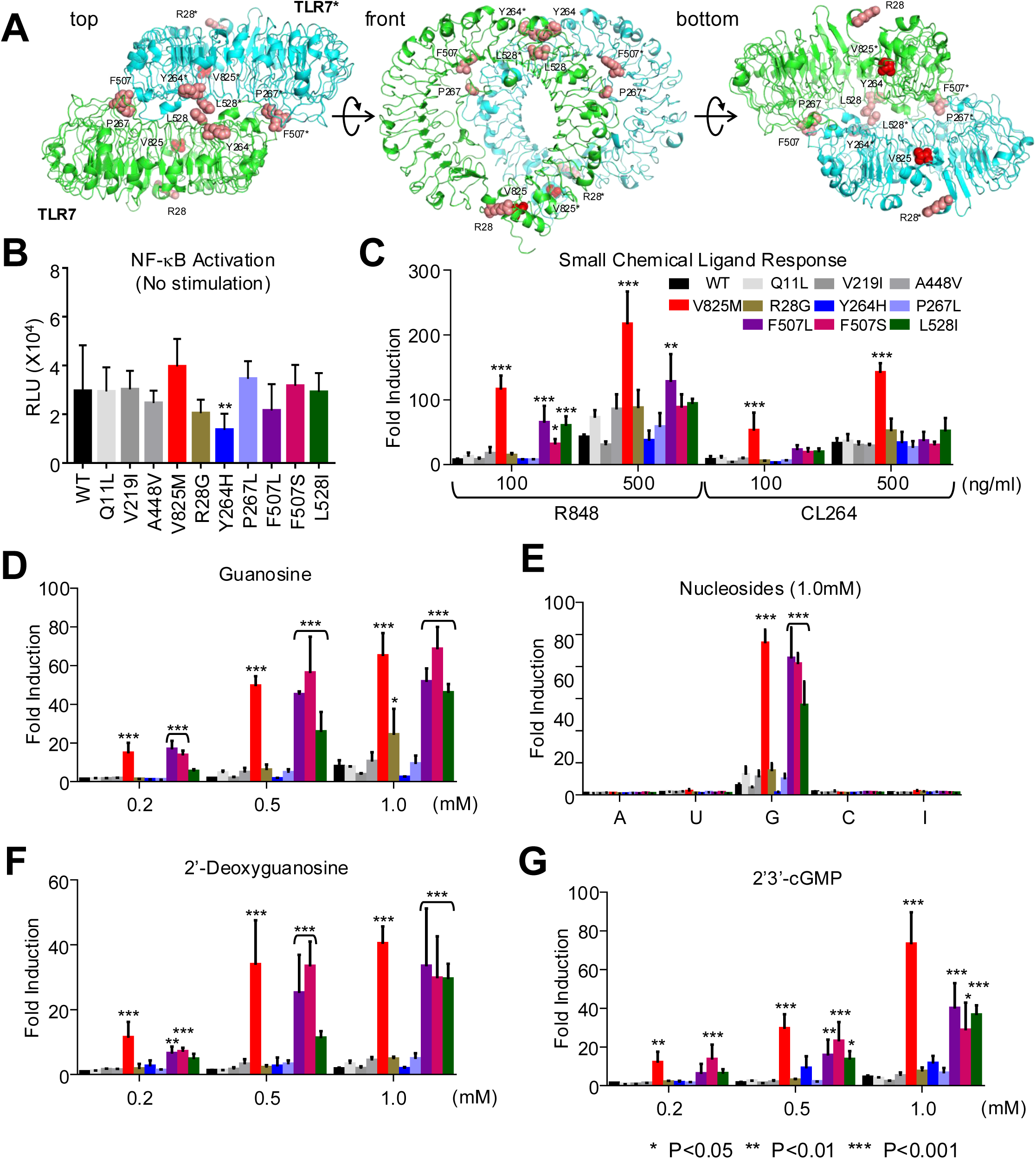
TLR7 V825M is a distinct gain-of-function variant robustly responding to a wide variety of TLR7 ligands. (**A**) Structure of TLR7 ectodomain and location of TLR7 V825M and known pathogenic variants. (**B-H**) NF-κB dependent luciferase reporter assay of human TLR7 variants in HEK 293T cells co-expressing TLR7 and UNC93B1. WT, wild type; V825M, variant identified in this study; R28G, Y264H, P267L, F507L, F507S, L528I, previously reported pathogenic variants; Q11L, V219I, A448V, benign variants from gnomAD. Cells were unstimulated (**B**), stimulated with R848/CL264 (**C**), guanosine (**D**), nucleosides (**E**), 2’-deoxyguanosine (dG) (**F**), 2’, 3’-cGMP (**G**). Data represent the mean fold induction of NF-κB activity ± SD, calculated as the RLU of stimulated cells divided by the RLU of non-stimulated cells. The data shown are representative of more than three independent experiments. *, p<0.05; **, p<0.01; ***, p<0.001

**Figure 5.**
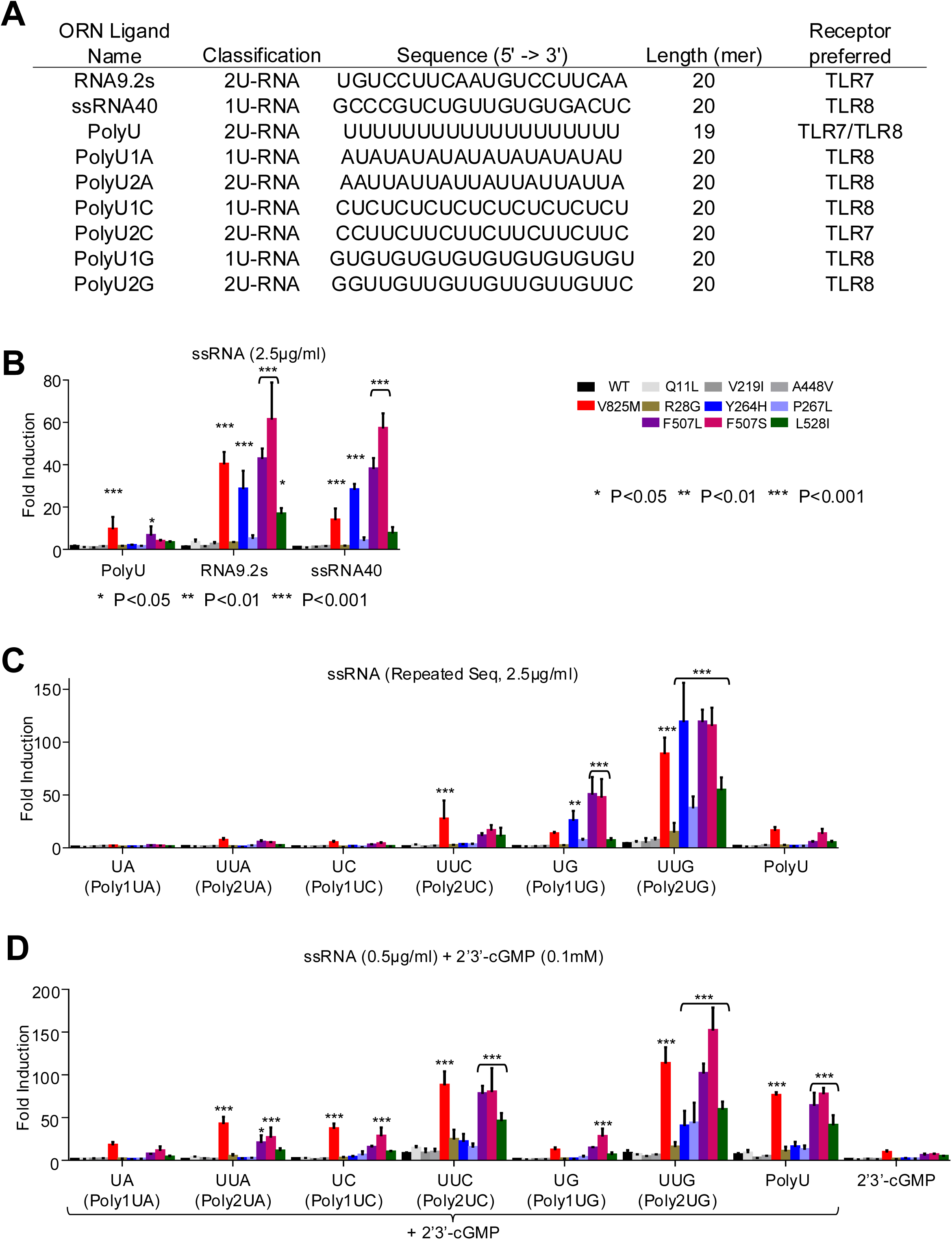
TLR7 V825M and a cluster of variants on dimerization surface relax ssRNA spectrum recognized and enhance synergic activation by 2’, 3’-cGMP and ssRNA. **(A)** Sequences of RNA ligands used in this study. **(B, C)** NF-κB dependent luciferase reporter assay of huTLR7 variants in HEK 293T cells co-expressing TLR7 and UNC93B1. WT, wild type; V825M, variant identified in this study; R28G, Y264H, P267L, F507L, F507S, L528I, previously reported pathogenic variants; Q11L, V291I, A448V, benign variants from gnomAD. Cells were stimulated with ssRNA ligand indicated (**B, C**) and 2’, 3’-cGMP with or without indicated ssRNA ligands (**D**).

## Discussion

In this study, we establish a new paradigm for TLR7-driven autoimmunity, in which not only the enhanced responses to canonical ligands but also the expanded ligand specificity by certain pathogenic *TLR7* genetic variant underlies the development of human lupus. We identified a novel *de novo TLR7* missense variant, V825M, in a patient with severe childhood lupus and demonstrated its pathogenicity through comprehensive functional assays and a murine model. Our findings show that specific genetic variations from monogenic lupus patients, including V825M, not only amplify TLR7 signaling in response to canonical ligands but, more critically, broaden the repertoire of ssRNA sequences. The creation of an orthologous knock-in mice (VarM mice) that spontaneously develops a lupus-like autoimmune disease confirms the pathogenic function of V825M *in vivo*. Notably, VarM mice recapitulate the patient’s duodenitis, a clinical feature not reported in other TLR7-dependent autoimmune models. This makes the VarM strain a valuable and unique tool for investigating how TLR7 contributes to intestinal immunity and pathology, distinct from existing SLE models.

Regarding the molecular mechanisms underlying hyperactivation of the V825M variant, we demonstrated that it is strictly ligand dependent. The V825M variant exhibited augmented responses to a wide range of canonical TLR7 ligands, including synthetic small-molecule agonists, guanosine analogs, and ssRNA ligands. Importantly, the V825M variant responded strongly to ssRNA sequences that are poor activators of WT TLR7, such as those with only a single uridine (1U-RNAs) like ssRNA40, a canonical TLR8 ligand. This observation provides a crucial insight into the specific drivers of lupus. For instance, the SLE-associated Y264H variant also enhanced responses to 1U-RNA ligands and causes lupus-like disease in mice, despite not conferring increased responsiveness to guanosine analogs. In contrast, SLC29A3 deficiency, which leads to the accumulation of guanosine analogs and excessive TLR7 activation (*25*), results in histiocytosis but not lupus. This discrepancy suggests that the acquisition of responsiveness to a broader spectrum of endogenous ssRNAs, particularly to 1U-RNA sequences, is a key determinant of lupus development in humans and mice.

The precise structural mechanism by which the LRR-CT variant V825M alters ligand specificity remains to be elucidated. However, our data provide important clues. Other lupus-associated variants located at the TLR7 dimerization interface (F507L, F507S, L528I) mimicked the expanded ligand recognition profile of V825M, suggesting the involvement of conformational changes in the dimeric structure of TLR7 for influencing ligand specificity.

Although V825 is not located at the direct dimerization interface, it is part of a conserved hydrophobic cluster proximal to a disulfide bond and the transmembrane domain. We speculate that the V825M substitution destabilizes this cluster and perturbs the dimerization interface. This could, in turn, reshape the architecture of the guanosine- and ORN-binding pockets, thereby lowering the activation threshold and broadening the spectrum of TLR7 ligands. Further investigation will be needed to address this hypothesis.

In conclusion, our study of a lupus patient carrying a novel *TLR7* variant provides the first genetic and molecular evidence that a change in ligand recognition specificity by an innate immune sensor is a primary cause of human lupus. This work redefines our understanding of how TLR7 gain-of-function variants can provoke autoimmunity, shifting the focus from simple hyperactivation to the critical role of an altered ligand spectrum.

## Material and methods

### Data reporting

No statistical methods were used to predetermine the sample size. The experiments were not randomized, and the investigators were not blinded to allocation during experiments, data analysis, and outcome assessment.

### Human subjects

All enrolled subjects provided written informed consent and were collected through protocols following local ethics and IRB approval at Chiba University Hospital (HS202207-05).

### Whole-exome sequencing and analysis

Genetic diagnosis was conducted via exome sequencing. In brief, genomic DNA was extracted from peripheral blood and enriched using the SureSelect XT Human All Exon V6 capture library (Agilent Technologies). Sequencing was performed on a HiSeq X Ten (Illumina) platform. Average depth was about 101X, and coverage of WES target regions with > 20X was ∼94.7%. Reads were aligned to the reference genome (hg19), and variants with a minor allele frequency above 5% in public databases or variants with fewer than five reads were excluded, except for known pathogenic variants cataloged in ClinVar and the Human Gene Mutation Database. We then focused on nonsynonymous single-nucleotide variants, insertions and deletions, and splice site variants.

### Human genomic DNA (gDNA) sequencing by Sanger dideoxy methods

The human gDNA was prepared from patients’ whole blood using a NucleoSpin® Blood L (U0954, Takara), following the indicated protocol. gDNA around *TLR7* exon 3 and SOX5 exon 5 were amplified by PCR using 50 ng of gDNA from the patient and controls. The primers used for the PCR were:

TLR7 exon3 forward primer; 5’- GCAAAAGAGAGGCAGCAAATGG -3’

TLR7 exon3 reverse primer; 5’- CAAAACACGCTTTTGGTGTG -3’

SOX5 isoform e exon5 forward primer; 5’-CTACACTACTTACCACCAAAGTAG-3’

SOX5 isoform e exon5 reverse primer; 5’-ATGTACTATCTATTGTGCCAGAGAG-3’.

These PCR amplicons were purified using DNA Clean & Concentrator (D4014, Zymo Research) and applied to Eurofins sequencing service (Eurofins Genomics) to obtain their sequences.

### Plasmid construction

Human TLR7 expression vector (pMX4 human TLR7 IRES rCD2), mouse Tlr7 expression vector (pMX4 mouse TLR7 IRES rCD2) and Unc93B1 expression vector (pMX4 human Unc93B1-EGFP CDS) were described previously (*4*). The pathogenic TLR7 variants (V825M, R28G, Y264H, F507L, P267V, F507S, L528I) and the Tlr7 variant (V826M) were generated by inverse PCR of the WT vectors. KOD-Plus-Mutagenesis Kit (SMK-101, Toyobo) was used to generate the *TLR7* and *Tlr7* variants. The benign TLR7 variants (Q11L, V219I, A448V) expression vectors (Myc DDK pCMV6 TLR7) were kindly provided by Jean-Laurent Casanova (The Rockefeller University) and coding sequences were subcloned into the pMX4 vector. All constructs were verified by Sanger sequencing, transformed into JM109 competent cells (9052, TaKaRa) for pMX vectors or DH5α (CC5204, Cosmo Bio) for Myc DDK pCMV vectors, amplified, and purified using NucleoBond® Xtra Midi/Maxi EF (U0420, Takara).

### Flow Cytometry

Spleens were minced with glass slides in RPMI 1640 medium, passed through a nylon mesh to remove debris, and treated with BD Pharm lysing buffer (BD Biosciences) to lyse red blood cells. Before surface staining, cells were incubated in a staining buffer (1× PBS with 2.5% FBS and 0.1% NaN₃) containing anti-mouse CD16/32 blocking mAb (clone 95). Subsequently, cells were stained for 15 mins on ice with fluorescent dye-conjugated mAbs against the following markers: CD11b (M1/70), FcγRIV (9E9), CD3ε (145-2c11), CD19 (6D5), CD11c (N418), CD71 (R17217), Ter119 (Ter119), Ly6C (HK1.4), and Ly6G (1A8). To detect endolysosomal TLR7, surface-stained cells were fixed and permeabilized using a Fixation/Permeabilization Solution Kit (BD Biosciences, USA) and stained again with PE-conjugated anti-mouse TLR7 mAb (clone A94B10). Stained cells were analyzed using an ID7000 Spectral Cell Analyzer (Sony Biotechnology). Flow cytometry data were processed using FlowJo v10.10.0 (BD Biosciences).

### Structural modeling of human TLR7 and UNC93B1

The structural model of Toll-like receptors in complex with UNC93B1 was provided by Protein Data Bank (PDB ID:7CYN). (*19*) TLR7 and UNC93B1 are shown in ribbon diagram.

### Cell lines and culture conditions

Human embryonic kidney–293T (American Type Culture Collection) cell lines were cultured in Dulbecco’s modified Eagle medium (DMEM)-high glucose (D5796, Sigma-Aldrich) complete medium supplemented with 10% (v/v) fetal bovine serum (FBS) (F7524, Sigma-Aldrich), 1% penicillin-streptomycin (15140122, Gibco) at 37°C and 5% CO_2_.

### Bone marrow-derived macrophages (BMDMs) culture and differentiation

Bone marrow cells were collected from the femur and tibia. Following erythrocyte lysis using an ACK Lysis Buffer [150mM ammonium chloride, 10mM potassium bicarbonate, 0.1mM EDTA], cells were filtered through a 70 µM mash and washed with DMEM-high glucose (D5796, Sigma-Aldrich) complete medium supplemented with 10% (v/v) fetal bovine serum (FBS) (F7524, Sigma-Aldrich), 1% penicillin-streptomycin (15140122, Gibco), 50µM 2-mercaptoethanol (21985023, Gibco). 1.0 × 10^7^ cells were seeded into Corning 10cm dish (430591, Corning) in full DMEM supplemented with 50 ng/ml macrophage colony-stimulating factor (315-02, Peprotech) and cultured at 5% CO2 at 37 °C. On day 5 and day 7, half of the medium was refreshed with full DMEM. On day 8, BMDMs were detached from the dish by adding TrypLE™ Express (12604013, Gibco) for 10 minutes at 37 °C and collected.

### Histopathology

For hematoxylin-eosin (H&E) staining and immunohistochemistry, spleen, kidney, liver, and lung tissues were fixed in 4% Paraformaldehyde phosphate-buffer solution (163-20145, FUJIFILM Wako Pure Chemical Corporation), and embedded in paraffin. 4-μm sections cut from formalin-fixed, paraffin-embedded (FFPE) tissues were stained with H&E or subjected to immunohistochemical staining with Mayer’s hematoxylin counterstaining. For liver immunohistochemistry, anti-F4/80 (1:500, D2S9R; rabbit monoclonal, CST) was used.

For immunofluorescence staining of kidney frozen sections, kidneys from WT and VarM mice were fixed in 4% paraformaldehyde, embedded in optimal cutting temperature (OCT) compound, snap-frozen, and stored at -80 °C until sectioning. 10 μm cryosections were Permeabilizeed in PBS containing 0.05% Triton X-100 (Nacalai tesque) and blocked with Blocking One Histo (Nacalai tesque) for 30 min. For immunostaining, Sections were incubated overnight at 4°C with FITC-conjugated anti-mouse Complement C3 antibody (55500, MP Biomedicals; 1:200 dilution), Alexa Fluor 594–conjugated goat anti-mouse IgG antibody (A11005, Thermo Fisher Scientific; 1:400 dilution), and DAPI (D1306, Thermo Fisher Scientific; 1:1000 dilution) diluted in Can Get Signal Immunostain Immunoreaction Enhancer Solution B (NKB-601, TOYOBO, Japan).

### NF-κB-dependent Luciferase Reporter Assay

The HEK 293T cells used for the reporter assay were cultured in RPMI 1640 medium (Nakalai tesque) supplemented with 10% FBS, 2 mM L-glutamine (Gibco), and 50 μM 2-mercaptoethanol. The cells were seeded at a density of 1 × 10⁶ cells/well in collagen-coated 6-well plates. To generate NF-κB reporter cells expressing human TLR7, HEK 293T cells in 6-well plates were transfected with 1 μg of human TLR7 cDNA in pMX-puro-IRES-rat CD2, 1 μg of WT human UNC93B1 cDNA in pMX-puro, and 25 ng of the pNL3.2.NF-κB-RE [NlucP/NF-κB-RE/Hygro] vector, using polyethyleneimine (PEI, "Max," MW 40,000; Polysciences, Inc.) and OPTI-MEM (Gibco). Twenty hours after transfection, the cells were re-seeded at approximately 5 × 10³ cells/well into collagen-coated 384-well white opaque plates (Corning). A few hours after re-seeding, the NF-κB reporter cells were stimulated with various ligands in the presence or absence of 15 μl/ml DOTAP (Roche) for 6 hours. Subsequently, Nanoluc activity was detected using the Nano-Glo Luciferase Assay System (Promega). Bioluminescence was measured as relative luminescence units (RLU)/second using the GloMax Explorer (Promega). To evaluate the activity of mouse Tlr7, the HEK 293T cells used for the reporter assay were cultured in DMEM-high glucose (D5796, Sigma-Aldrich) complete medium supplemented with 10% (v/v) FBS, 1% penicillin-streptomycin (15140122, Gibco). The cells were seeded at a density of 0.7 × 10^5^ cells/well in collagen-coated 24-well plates. To generate NF-κB reporter cells expressing mouse TLR7, HEK 293T cells in 24-well plates were transfected with 50 ng of mouse TLR7 cDNA in pMX-puro-IRES-rat CD2, 100 ng of WT human UNC93B1 cDNA in pMX-puro, 30 pg of the Renilla luciferase reporter plasmid (pRL-TK; E2241, Promega), and 100 ng of the firefly luciferase plasmid driven by the NF-κB response element vector (pGL4.32-luc2P/NF-κB-RE/Hygro; E8491, Promega), using polyethyleneimine and OPTI-MEM. Twenty hours after transfection, the NF-κB reporter cells were stimulated with indicated TLR7 ligands for 6 hours. Subsequently, Nanoluc activity was detected using the Dual-Luciferase® Reporter Assay System (E1910, Promega). Bioluminescence was measured as relative luminescence units (RLU)/second using a GloMax® 20/20 Luminometer. R848 and CL264 were purchased from InvivoGen (Hong Kong). Guanosine and 2′ -deoxyguanosine hydrate were purchased from MP Biomedicals (Santa Ana, CA, USA). Guanosine- 2’, 3’- cyclic monophosphate (2’,3’-cGMP) was purchased from BIOLOG Life Science Institute (Bremen, Germany). 20-mer phosphorothioate oligoribonucleotides, including RNA9.2s, ssRNA40, PolyU, PolyU1A, PolyU2A, PolyU1C, PolyU2C, PoluU1G, and PolyU2G, were synthesized by FASMAC (Kanagawa, Japan).

### Western blot analysis

For Western blot analysis, HEK 293T and BMDM were washed in ice-cooled PBS, scraped, and lysed in M-RIPA buffer [50 mM HEPES, 150 mM NaCl, 1 mM EDTA, 10% glycerol, 0.5% Sodium deoxycholate, 0.1% SDS, Halt™ Phosphatase Inhibitor Cocktail (78420, Thermo Fisher Scientific), protease inhibitor cocktail (P8340, Sigma Aldrich)]. After a 30-minute incubation on ice, the lysates were centrifuged at 15000g for 15min at 4℃, and the debris was removed. Lysates were denatured in SDS-polyacrylamide gel electrophoresis (PAGE) buffer [NuPAGE™ LDS Sample Buffer (4X) (NP0008, Invitrogen) + 5% of 2-mercaptoethanol (DTT)] at 75°C for 10 minutes. Proteins were separated by SDS-PAGE (Tris-Acetate, 1.5 mm, Mini Protein Gels, Invitrogen) and transferred to 0.45µm Nitrocellulose Membranes (Bio-Rad) in a Trans-Blot Turbo transfer system (Bio-Rad). Membranes were probed with the indicated antibodies and developed using the SuperSignal™ West Pico PLUS Chemiluminescent Substrate (34580, Thermo Fisher Scientific) and Bio-Rad ChemiDoc imaging system.

The following antibodies were used for Western blots: anti-mouse-Tlr7 (1:1000, #82658, Cell Signaling), anti-Phospho-IKKα/IKKβ (1:1000, C84E11, 2078, Cell Signaling), anti-IKKβ (1:1000, D30C6, 8943, Cell Signaling), anti-Phospho-IκBα (1:1000, EPR3148, ab92700, abcam), anti-IκBα (1:1000, L35A5, 4814, Cell Signaling), Secondary antibodies: Amersham ECL Mouse IgG, HRP-linked F(ab’)₂ fragment from sheep (1:5000, NA9310, Cytiva), HRP-conjugated Goat Anti-Rabbit IgG (H+L) (1:5000, SA00001-2, Proteintech).

### Detection of ANAs

ANAs were determined using a Kallestad HEp-2 Kit (60030, Bio-Rad). Briefly, serum was diluted at 1:40 and incubated on Hep-2 slides according to the manufacturer’s instructions. Since the kit was for human use, a goat anti-mouse IgG(H+L) secondary antibody FITC (#31569, Invitrogen) was used as a secondary antibody. The slides were imaged using the EVOS M7000 Imaging System (Thermo Fisher Scientific).

### Enzyme-Linked Immunosorbent Assay (ELISA)

Mouse bone marrow-derived macrophages (BMDM) were cultured in flat-bottom 96-well plates (BD Falcon, USA) at a density of 5 × 10^4^ cells per well. BMDM were stimulated with the indicated ligands (R848; 1nM, Poly U; 5µg/mL, RNA9.2s; 5µg/mL, Guanosine; 100µM, 2’-Deoxyguanosine, 2′, 3′-cGMP; 200µM, CpG; 100nM, Lipid A; 100ng/mL) for 16–20 hours, and IL-12p40 concentrations in the culture supernatants were measured using a Ready-Set-Go! ELISA kit (eBioscience, USA) according to the manufacturer’s instructions.

For R848 and LPS stimulation, BMDMs were seeded at 0.2×10^6^ cells per well into tissue culture-treated flat-bottom 12-well plates. The next day, cells were stimulated with several concentrations of LPS or R848 for 8 hours, and the supernatant was collected. TNF-α and IL-6 concentrations were measured using Solid Phase Sandwich ELISA kits (DY410 and DY406, R&D Systems) according to the manufacturer’s instructions.

### Animals

C57BL/6JJcl (B6) and B6C3F1/Jcl (B6C3F1) were purchased from CLEA Japan Inc (Tokyo, Japan). All mice were maintained under the following conditions: 23 ± 2 ℃, 50% ± 20% humidity, 12:12 h dark light period in micro isolator cages under specific pathogen-free conditions. Food and water were supplied ad libitum. All animal experiments to generate the gene knock-in mice were approved by the Institutional Animal Care and Use Committee at NIG or the Animal Committee of the Graduate School of Medicine, Chiba University (approval identification A6-250).

### Preparation of genome editing mixture

HiFi Cas9 protein was purchased from IDT (Alt-R™ S.p. HiFi Cas9 Nuclease V3, Integrated DNA Technologies, IA, USA). The guide RNA (gRNA) (Rev:CTCACACGTATACAGATCAA) was designed using CRISPOR (http://crispor.gi.ucsc.edu/) and Custom Alt-R™ CRISPR-Cas9 guide RNA (https://sg.idtdna.com/site/order/designtool/index/CRISPR_CUSTOM) and formed by annealing crRNA and tracrRNA purchased from IDT. The ssODN (TGTGACTTGTGTAGGTCCAGGAGCACACAAAGGTCAAAGTATGATATCGCTTGATCTGTATACGTGTGAGTTAGATCTCACAAACCTGATTCT) was synthesized by Eurofins Genomics (Tokyo, Japan) and used for HDR donor template. The final concentrations of the genome editing mixture were 100 ng/µl (Cas9 protein), 70 ng/µl (gRNA), and 300 ng/µl (ssODN).

### Electroporation of mouse zygotes

B6 female mice were superovulated by intraperitoneal (IP) injection of 100 µl CARD HyperOva (Kyudo Co., Ltd., Saga, Japan) followed 48 h later by IP injection of 5 IU human chorionic gonadotropin (Gonatropin, ASKA Animal Health Co., Ltd. Tokyo, Japan). Seventeen hours after the administration of human chorionic gonadotropin, egg cells were collected from superovulated female mice and inseminated with pre-incubated spermatozoa of B6 male mice in HTF medium (ARK Resource Co., Ltd. Kumamoto, Japan). Zygotes were washed to remove cumulus cells and transferred into the KSOM medium (ARK Resource). Electroporation was performed using the electrode (CUY501P1-1.5, Nepa Gene, Chiba, Japan) and the electroporator (NEPA21, Nepa Gene). Zygotes were placed in the electrode gap filled with 4 µl of genome editing mixture diluted in Opti-MEM (Thermo Fisher Scientific, Waltham, MA, USA). Electroporation was conducted using the following parameters: Poring Pulse: 40 V, 3.5-ms pulse width, 50-ms pulse interval, 4 pulses, 10% decay, + polarity, and Transfer Pulse: 7 V, 50-ms pulse width, 50-ms pulse interval, 5 pulses, 40% decay, ± polarity. After electroporation, zygotes were returned to the KSOM medium and incubated until the 2-cell stage under 5% CO2 at 37 ℃. All surviving 2-cell embryos were transferred into the oviduct of pseudopregnant B6C3F1 females anesthetized with 2-4% isoflurane (Viatris Inc, PA, USA).

### Mouse genotyping analysis

Genomic DNA from each animal was extracted from ear biopsy samples or tail-tip and used for PCR to detect the *Tlr7*-V826M variant. The targeted region was amplified with a pair of flanking primers: Tlr7-Typing-F (TGCGCTATCTAGACATCAGTTC) and Tlr7-Typing-R (ATCTTCCAATTTTGCCACCAGC). The PCR reaction was performed using Tks Gflex™ DNA Polymerase (Takara Bio Inc, Shiga, Japan). These PCR amplicons were applied to Eurofins sequencing service (Eurofins Genomics) to obtain their sequences.

### Statistical Analyses

Data were analyzed using GraphPad Prism 8.0. Depending on experimental design, statistical significance was tested via either two-tailed unpaired or paired Student’s t-test or one-way ANOVA.

## Data Availability

Supplementary Text is available from the corresponding author upon reasonable request.

## Author contributions

All authors read and provided feedback on the submitted manuscript.

Conceptualization: KaM

Investigation: AT, KaM, TaS, ST, FY, YG, KS, KE, TaK, TaI, AM, YuN, HY, TY, JI

Variant identification and interpretation: YoN, TaO, KaM, ToI

Structural analysis and visualization: ToS, TN

Generation of knock-in-mice: YI, TsK

Funding acquisition: KaM, TaS, KeM, HN, ToO

Supervision: KeM, HN, KH, HH, ToO, EN

Writing – original draft: KaM, TaS, AT

Writing – review & editing: ToS, KeM, HN

## Acknowledgments

We thank the patient, family members, and healthy volunteers for participating in this study; Jean-Laurent Casanova, Bertrand Boisson, and Takaki Asano for providing the MycDDK pCMV6 TLR7 expression vectors and their information (WT and benign variants); Emi Utsuno for genetic counseling to the family; and Maho Yoshino, Yuzuho Nakagawa for technical assistance.

---

**Figure S1.**
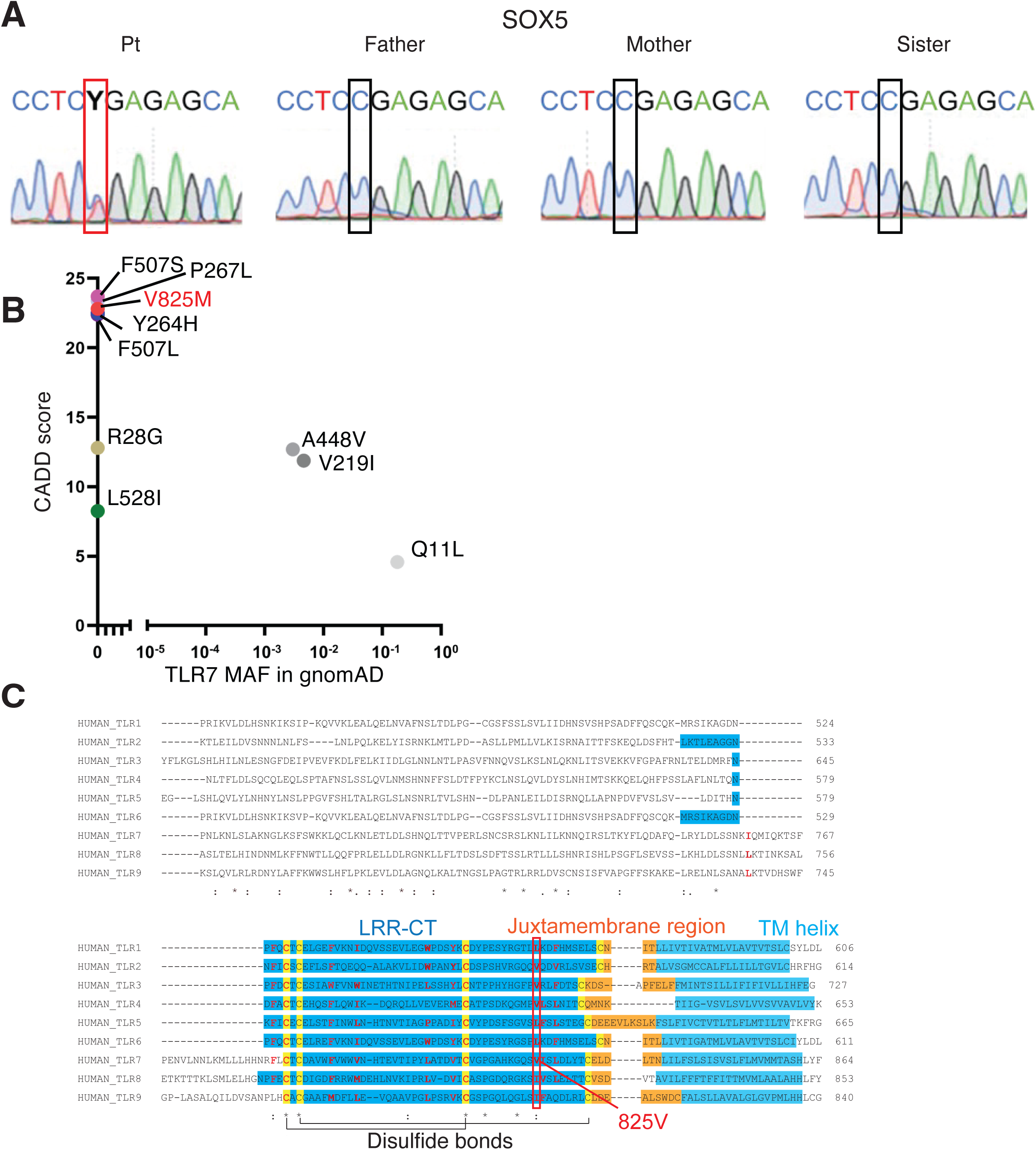
A novel missense variant in the LRR-CT domain of TLR7 associated with severe childhood lupus. (**A**) Chromatograms of Sanger sequencing of gDNA from the patient and family for SOX5 R203X variant. Y indicates C or T. (**B**) Population genetics of TLR7 variants used in this study. CADD, combined annotation-dependent depletion; MAF, minor allele frequency. (**C**) Sequence alignment around LRR-CT (blue), juxtamembrane region (orange), and TM helix (light blue) among human TLRs. LRR-CT, leucine-rich repeat-C terminal; red, amino acids forming hydrophobic core; yellow, disulfide bonds; red box, conserved hydrophobic amino acids corresponding to TLR7 825V.

**Figure S2.**
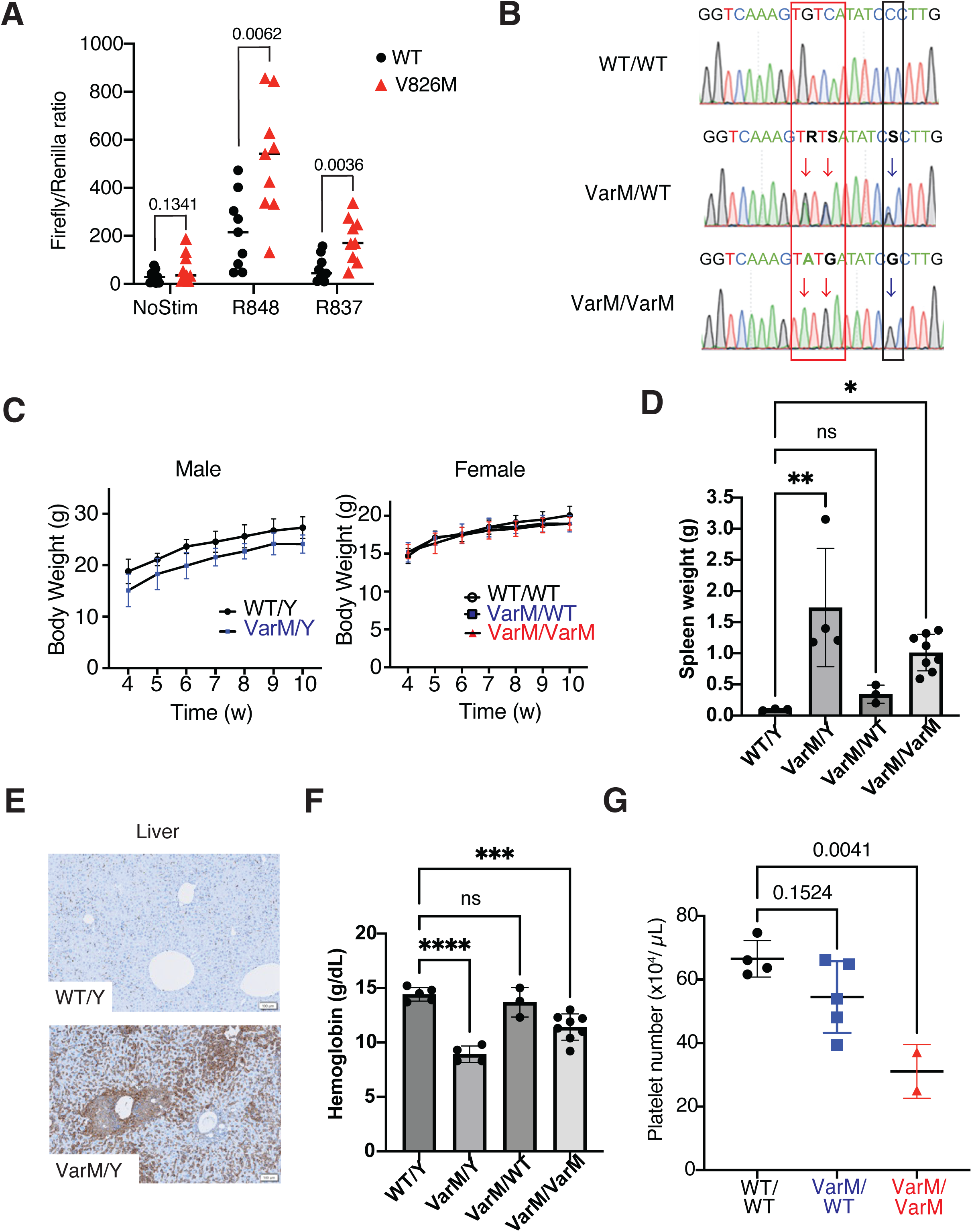
VarM mice develop lupus-like autoimmune disease. (**A**) NF-κB dependent luciferase reporter assay of mouse Tlr7 variant V826M in HEK 293T cells co-expressing UNC93B1. Data indicate the mean of nine biological replicates. Statistical analysis was performed using unpaired t-test, and exact *p* values are indicated. (**B**) Chromatograms of Sanger sequencing of gDNA for Tlr7 V826M. Red arrow, base changes for amino acid alternation; blue arrow, synonymous base change for ssODN protection from Cas9 digestion. R, A or G; S, C or G. (**C**) Body weight of male mice (left, WT/Y and VarM/Y, n=6 for each genotype) and female mice (right, WT/WT, VarM/WT, and VarM/VarM, n=6 for each genotype). Data indicate mean ± SD. (**D**) Weight of spleen from 52-week-old mice with indicated genotypes. Data indicate mean, and statistical analyses were performed by one-way ANOVA. *, p<0.05; **, p<0.01; NS, not significant. **(E**) Immunofluorescence staining showing infiltration of F4/80^+^ cells in the liver in a 52-week-old male VarM mouse. (**F**) Hemoglobin levels of indicated mice at the age of 52 weeks. Data indicate the average of n=3 (WT/Y), n=4 (VarM/Y), n=3 (VarM/WT), n=8 (VarM/VarM) mice. Statistical analysis was performed using one-way ANOVA, Data indicate mean, and statistical analyses were performed by one-way ANOVA. *, p<0.05; **, p<0.01; NS, not significant. (**G**) Platelet count in peripheral blood from indicated mice at the age of 20 weeks. Data indicate mean of n=4 (WT/WT), n=5 (VarM/WT), and n=2 (VarM/VarM). Statistical analyses were performed using one-way ANOVA. Data indicate mean, and statistical analyses were performed by one-way ANOVA. *, p<0.05; **, p<0.01; NS, not significant.

**Figure S3.**
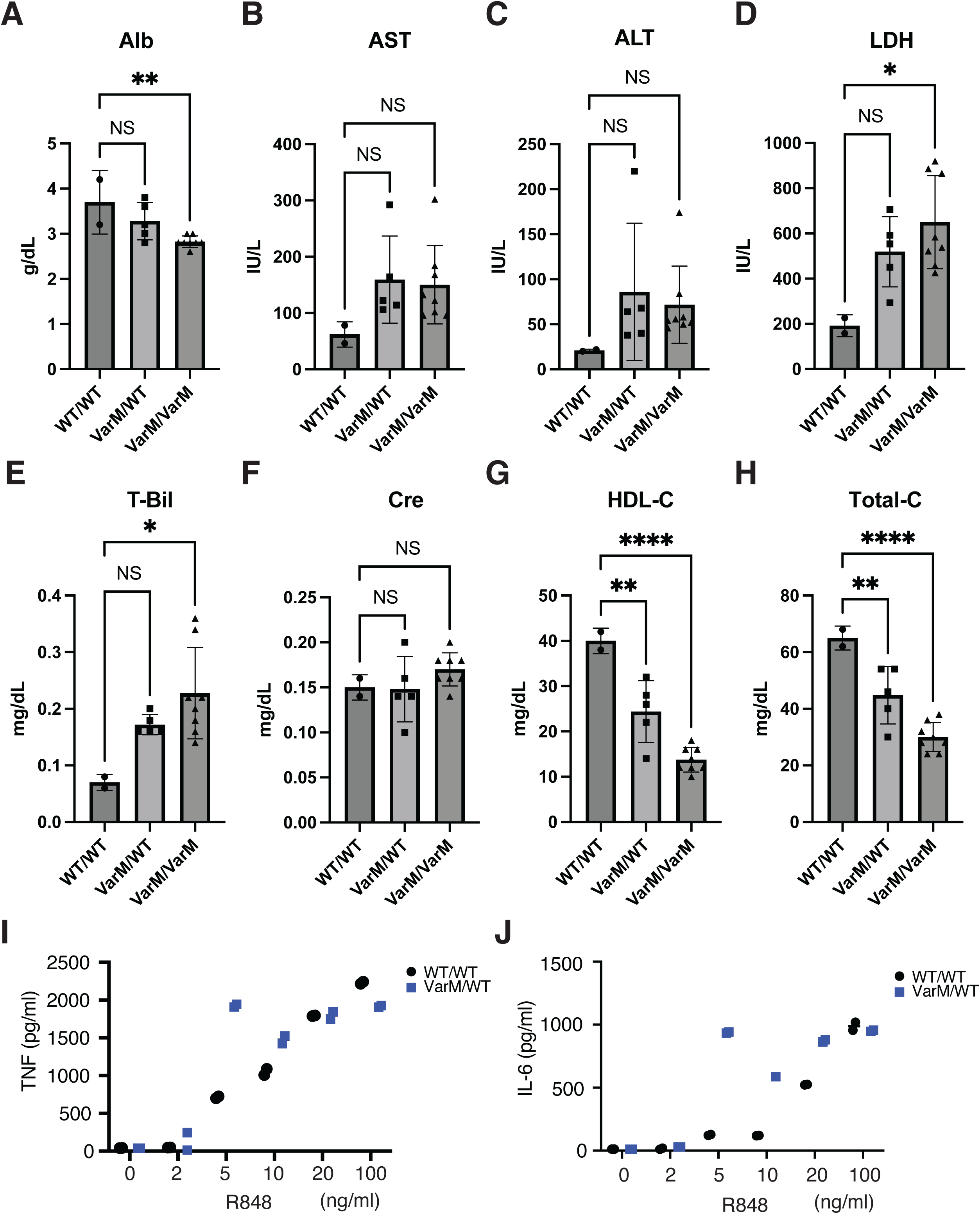
(**A-H)** Biochemical analysis of serum from VarM mice. Alb, albumin; AST, aspartate aminotransferase; ALT, alanine aminotransferase; LDH, lactate dehydrogenase; T-Bil, total bilirubin; Cre, creatinine; HDL-C, high-density lipoprotein cholesterol; Total-C, total cholesterol. Statistical analysis was performed using one-way ANOVA. *, p<0.05; **, p<0.01; ***, p<0.001; NS, not significant. (**I, J**) Pro-inflammatory cytokine TNF (**I**) and IL-6 (**J**) production in BMDM from WT/WT or VarM/WT mice at the age of 10 weeks. Data indicate an average of n=2 (WT/WT), n=2 (VarM/WT).

## Supplementary Text

Supplementary Text is available from the corresponding author upon reasonable request.

